# Changes in urinary glutathione sulfonamide (GSA) levels between admission and discharge of patients with cystic fibrosis

**DOI:** 10.1101/2023.10.24.23297497

**Authors:** Tamara L Blake, Peter D Sly, Isabella Andersen, Claire E Wainwright, David W Reid, Scott C Bell, Anthony J Kettle, Nina Dickerhof

## Abstract

There is an urgent need to develop sensitive, non-invasive biomarkers that can track airway inflammatory activity for patients with cystic fibrosis (CF). Urinary GSA levels correlate well with GSA levels in BAL samples and other markers of neutrophilic inflammation, suggesting that this biomarker may be suitable for tracking disease activity in this population.

We recruited 102 children (median 11.5 years-old) and 64 adults (median 32.5 years-old) who were admitted to hospital for management of an acute pulmonary exacerbation and/or eradication of infectious agents such as *P. aeruginosa* or *S. aureus*. Our aim was to explore how urinary GSA levels changed across admission timepoints. Urine samples were collected at admission and discharge, and GSA measured by liquid chromatography with mass spectrometry. Paired admission-discharge results were compared using Wilcoxon signed-rank test.

Paired admission-discharge samples were available for 49 children and 60 adults. A statistically significant difference was observed between admission-discharge for children, but not for adults. Spearman’s correlation analysis identified a correlation between urinary GSA levels and age, sex, inflammatory markers, and *P. aeruginosa* infection for children only. Our preliminary findings suggest that urinary GSA is responsive to the resolution of an acute pulmonary exacerbation and therefore warrants further studies in this population.

---

Data from the Australian Early Respiratory Surveillance Team for Cystic Fibrosis (AREST CF) program suggests that pulmonary inflammation, infection, and structural disease can be present not only in the absence of clinical symptoms, but in patients as young as 3 months of age.(1,2) Furthermore, acute pulmonary exacerbations, particularly those requiring hospitalisation, are significantly correlated with loss of lung function and adverse lung health outcomes.(3,4) Published data suggest that exaggerated neutrophilic inflammation is a primary driver of early CF lung disease progression.(1,2,5–7) Activated neutrophils release myeloperoxidase, which catalyses the formation of hypochlorous acid (HOCl), resulting in tissue damage. Glutathione (GSH), a normally plentiful antioxidant in the airways, scavenges HOCl and is oxidized mainly to glutathione disulfide (GSSG) and mixed protein disulfides.(5,7) Glutathione sulfonamide (GSA) is also formed as a minor but HOCl-specific and irreversible oxidation product, making it an attractive biomarker of neutrophil-derived oxidative stress. Elevated levels of GSA in bronchoalveolar lavage (BAL) and serum samples from young children with CF correlate with *P. aeruginosa* infections and radiological bronchiectasis.(8) However, routine collection of BAL samples is challenging, especially in children. There is an urgent need to explore and develop sensitive, non-invasive biomarkers for use in this population. Urinary GSA levels have been shown to correlate well with GSA levels in BAL samples and other markers of neutrophilic inflammation,(8) suggesting that urinary GSA may be suitable for identifying and tracking inflammatory disease activity for patients with CF.

We recruited 102 children (median age 11.5 years, 25%-75% 6.4-14.4) and 64 adults (median age 32.5 years, 25%-75% 25.0-39.0) who were admitted to hospital for management of an acute pulmonary exacerbation and/or eradication of infectious agents such as *P. aeruginosa* or *S. aureus*. All participants reported an increase in wet cough and/or sputum production at the time of their admission. Our aim was to explore how urinary GSA levels varied between admission and discharge. Clinically relevant information – height, weight, markers of systemic inflammation (from BAL and blood samples), microbiology results (sputum samples), spirometry results, self-reported symptoms – where collated from medical records. Written informed consent was obtained from all parents/guardians or participants if >18 years of age. Ethics was approved by the Children’s Health Queensland HREC.

GSA was measured by liquid chromatography with mass spectrometry (LC-MS) using multiple reaction monitoring on a Sciex 4000 QTrap as previously described (8) however, normalization using specific gravity was not performed (account for urine dilution). GSA results were reported as medians with 25^th^-75^th^ percentiles. Differences in urinary GSA results at collection timepoints were compared using Kruskal-Wallis rank test followed by Dunn’s method for pairwise multiple comparisons. Paired admission and discharge urinary GSA results were analysed using Wilcoxon signed-rank test. Data from children and adults were analysed separately as inflammatory biomarkers vary with disease stage.(9,10)

For children, 122 admissions were documented during the study period with a total of 145 samples collected: 87 on day 1 of admission and 58 on day of discharge. For adults, 64 admissions were documented with a total of 124 samples collected: 64 on day 1 of admission and 60 on day of discharge. For children, the mean length of admission was 14 days (SD 4, range 7-39 days) and for adults, the mean length of admission was 9 days (SD 3, range 6-19 days). Significant differences were observed between children and adults for age, height, weight, inflammatory markers (neutrophil elastase [NE] activity, interleukin 8 [IL-8], matrix metallopeptidase 9 [MMP-9], myeloperoxidase [MPO], high sensitivity c-reactive protein [hsCRP]), and *P. aeruginosa* and *S. aureus* infection history variables (Table 1).

**Table 1:**
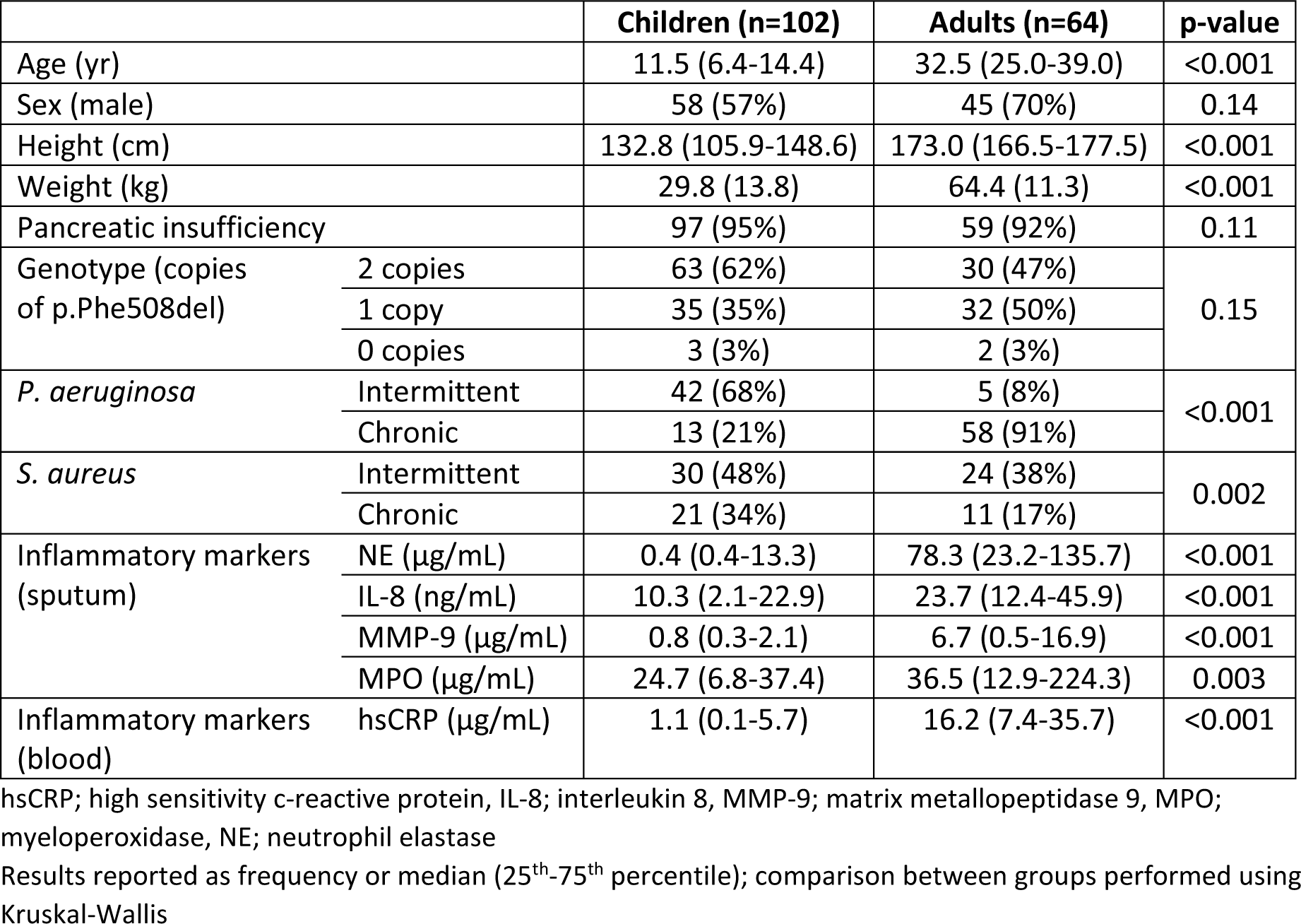
Demographics and clinical characteristics on admission.

|  |  | Children (n=102) | Adults (n=64) | p-value |
| --- | --- | --- | --- | --- |
| Age (yr) |  | 11.5 (6.4-14.4) | 32.5 (25.0-39.0) | <0.001 |
| Sex (male) |  | 58 (57%) | 45 (70%) | 0.14 |
| Height (cm) |  | 132.8 (105.9-148.6) | 173.0 (166.5-177.5) | <0.001 |
| Weight (kg) |  | 29.8 (13.8) | 64.4 (11.3) | <0.001 |
| Pancreatic insufficiency |  | 97 (95%) | 59 (92%) | 0.11 |
| Genotype (copies of p.Phe508del) | 2 copies | 63 (62%) | 30 (47%) | 0.15 |
|  | 1 copy | 35 (35%) | 32 (50%) |  |
|  | 0 copies | 3 (3%) | 2 (3%) |  |
| *P. aeruginosa* | Intermittent | 42 (68%) | 5 (8%) | <0.001 |
|  | Chronic | 13 (21%) | 58 (91%) |  |
| *S. aureus* | Intermittent | 30 (48%) | 24 (38%) | 0.002 |
|  | Chronic | 21 (34%) | 11 (17%) |  |
| Inflammatory markers (sputum) | NE (µg/mL) | 0.4 (0.4-13.3) | 78.3 (23.2-135.7) | <0.001 |
|  | IL-8 (ng/mL) | 10.3 (2.1-22.9) | 23.7 (12.4-45.9) | <0.001 |
|  | MMP-9 (µg/mL) | 0.8 (0.3-2.1) | 6.7 (0.5-16.9) | <0.001 |
|  | MPO (µg/mL) | 24.7 (6.8-37.4) | 36.5 (12.9-224.3) | 0.003 |
| Inflammatory markers (blood) | hsCRP (µg/mL) | 1.1 (0.1-5.7) | 16.2 (7.4-35.7) | <0.001 |

Results reported as frequency or median (25<sup>th</sup>-75<sup>th</sup> percentile); comparison between groups performed using Kruskal-Wallis

Matched admission-discharge samples were available for 49 children and 60 adults. For children, a statistically significant difference was observed between admission and discharge results (0.15 [0.07-0.20] μM vs. 0.09 [0.06-0.14] μM, p=0.024). Thirty-four children (69.3%) had lower GSA results at discharge (compared to admission), while 15 (30.6%) had an increase in GSA results at discharge (Figure 1, left panel). For adults, no statistically significant difference was observed between admission and discharge results (0.05 [0.03-0.09] μM vs. 0.05 [0.03-0.07] μM, p=0.078). Thirty-six adults (60%) had lower GSA results at discharge (compared to admission), while 24 (40%) had higher GSA results on discharge (Figure 1, right panel).

**Figure 1:**
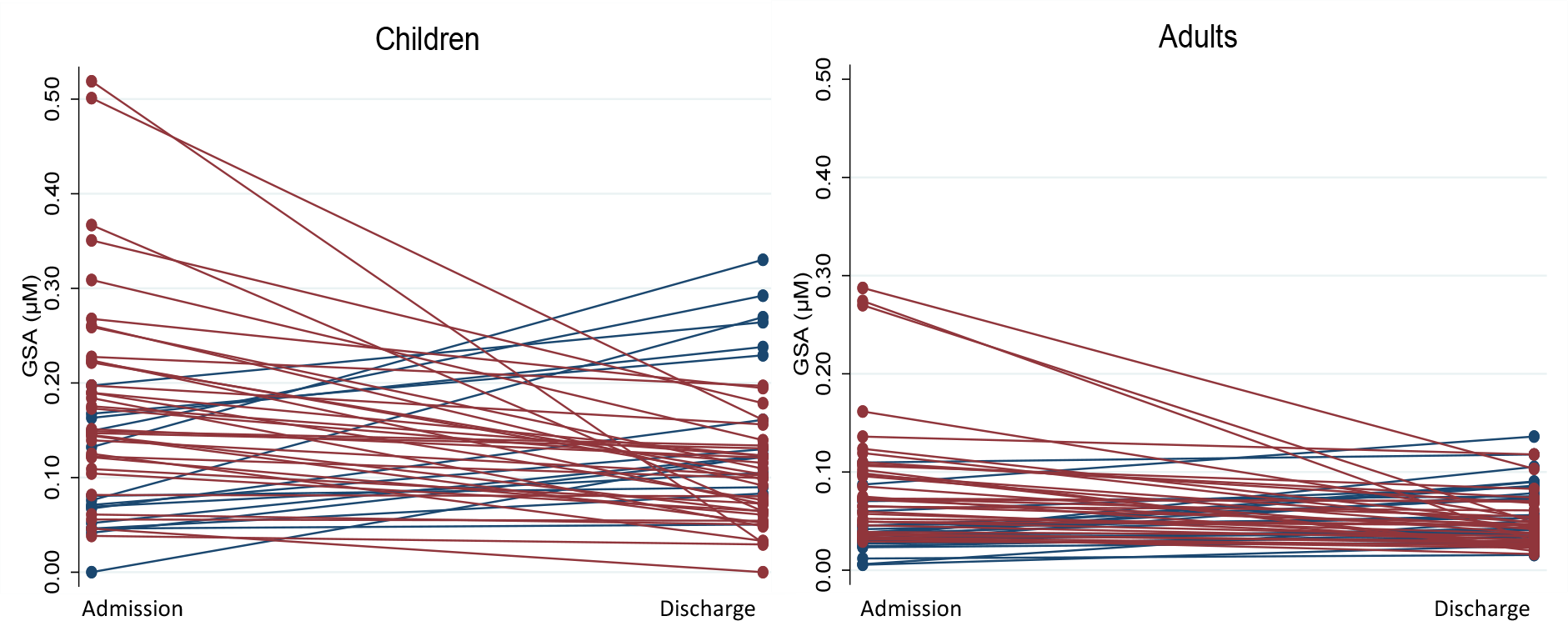
Matched admission-discharge urinary GSA results for children (left panel) and adults (right panel) with CF. Red lines indicate patients who had higher GSA values at admission compared to discharge (69.3% for children and 60% for adults). Blue lines indicate patients who had higher GSA values at discharge compared to admission (30.6% for children and 40% for adults).

We performed Spearman’s correlation analysis on GSA results collected at the time of admission with age, height, weight, lung function, infection status (absence vs. presence), and inflammatory markers. Analysis was performed separately for children and adults. No associations were seen between any variables and GSA values for adults. For children, a weak positive correlation was identified between urinary GSA levels and age (Rho=0.113, p=0.04), sex (Rho=0.110, p=0.04), NE activity (Rho=0.168, p=0.01) and MMP-9 (Rho=0.175, p=0.01). A weak negative correlation was also seen between urinary GSA and *P. aeruginosa* infection (Rho=-0.310, p=0.02) however, this may be due to data coding (presence vs. absence rather than by a quantifiable measure of infection burden such as colony forming units).

In this pilot study, higher levels of urinary GSA were seen at the beginning of an admission compared to those at discharge for children however, no significant differences were observed in adults. It is important to note that our samples were not corrected for urine dilution, as such, results may be impacted by dehydration at the time of collection. All participants were symptomatic at the time of their admission with an increase in wet cough and/or sputum production. Upon discharge, both children and adult participants showed significant improvement in forced expiratory volume in 1 second (FEV_1_) z-score values, and significantly reduced markers of systemic inflammation (including NE activity, MMP-9 and hsCRP). Reduction of GSA levels following treatment of an exacerbation in children provides supporting evidence to urinary GSA as a potential biomarker of pulmonary neutrophil-derived oxidative stress that can be used to non-invasively monitor changes in pulmonary inflammation activity. The lack of significant findings in adults requires additional studies, but may be due to compromised antioxidant defences contributed to by diminishing GSH levels in epithelial lining fluid as patients age.(11)

Many traditional measures of disease activity – lung function, serum analysis, sputum/BAL samples – are not feasible in young patients or are unrealistic to repeatedly perform. For example, spirometry cannot reliably be performed in patients ≤3 years of age(12) and other alternatives, such as the multiple breath washout, require sedation in very young patients.(13) Other objective measures including increased cough, changes in sputum volume or consistency, decreased appetite, weight loss and increased lethargy(14,15) have been used. These measures, however, can vary significantly between patients experiencing an exacerbation. Techniques that are sensitive to changes in disease activity, can easily be performed on patients of all ages, and can quickly return accurate results in routine clinical settings are needed to identify patients at risk of future exacerbations to allow for more timely and targeted intervention strategies.

Our preliminary findings suggest that urinary GSA is responsive to the resolution of an acute pulmonary exacerbation and correlates with other measures of disease activity for children with CF. In adults experiencing an exacerbation, urinary GSA were not as informative but the lower levels in adults compared to children suggest biological differences that need further investigation. This is a relatively small sample of opportunistically collected data largely pertaining to patients admitted with symptoms of an acute pulmonary exacerbation. Larger prospective cohort studies including participants <6 years of age, that routinely measure urinary GSA during both stable and unwell periods, alongside other measures of clinical status are needed to further explore the potential of this biomarker in the CF population. In addition, future studies should measure urine specific gravity to account for dehydration as a confounding factor.

## Data Availability

All data produced in the present study are available upon reasonable request to the authors.

## Acknowledgements

The study team would like to acknowledge all patients and families who contributed to this work. We especially thank the CF clinical nursing team at the Queensland Children’s and Prince Charles Hospitals for their assistance with coordinating sample collection.

## Funding Sources

This work has been supported by the Cystic Fibrosis Foundation (USA, SLY18KO) and Children’s Hospital Foundation (AUS).

## Conflict of Interest Statement

T.L. Blake holds a Children’s Hospital Foundation ECR Fellowship for salary support and an Australian CFA Research Trust Innovation Grant for unrelated research. P.D. Sly holds NHMRC Investigator and Clinical Trials & Cohort Study grants. Authors C.E. Wainwright and S.C. Bell have received institutional payments for chairing and speaking at educational events (Vertex Pharmaceuticals). C.E. Wainwright also declares participating in steering committee and advisory board sessions for Vertex Pharmaceuticals. N. Dickerhof holds a Sir Charles Hercus Research Fellowship from the Health Research Council of New Zealand. Remaining authors I. Andersen, D.W. Reid and A.J. Kettle declare that they have no conflicts of interest.

## Credit Roles

**Tamara Blake:** Conceptualization, Formal Analysis, Visualization, Writing – original draft, review & editing; **Peter Sly:** Conceptualization, Methodology, Visualization, Writing – review & editing; **Isabella Andersen:** Investigation, Resources, Writing – review & editing; **Claire Wainwright:** Conceptualization, Methodology, Writing – review & editing; **David Reid:** Conceptualization, Methodology, Writing – review & editing; **Scott Bell:** Conceptualization, Methodology; Writing – review & editing, **Anthony Kettle:** Conceptualization, Methodology, Investigation, Writing – review & editing; **Nina Dickerhof:** Conceptualization, Methodology, Investigation, Writing – review & editing

## Notes

### Competing Interest Statement

TL Blake holds a Childrens Hospital Foundation ECR Fellowship for salary support and an Australian CFA Research Trust Innovation Grant for unrelated research. PD Sly holds NHMRC Investigator and Clinical Trials & Cohort Study grants. Authors CE Wainwright and SC Bell have received institutional payments for chairing and speaking at educational events (Vertex Pharmaceuticals). CE Wainwright also declares participating in steering committee and advisory board sessions for Vertex Pharmaceuticals. N Dickerhof holds a Sir Charles Hercus Research Fellowship from the Health Research Council of New Zealand. Remaining authors Andersen, Reid and Kettle declare that they have no conflicts of interest.

### Funding Statement

This study was funded by the Cystic Fibrosis Foundation (USA, SLY18KO) and Children's Hospital Foundation (AUS).

### Author Declarations

Children's Health Queensland Hospital and Health Service Human Research Ethics Committee gave ethical approval for this work.

## References

1. Sly PD, Brennan S, Gangell C, de Klerk N, Murray C, Mott L, et al. Lung disease at diagnosis in infants with cystic fibrosis detected by newborn screening. Am J Respir Crit Care Med. 2009 Jul 15;180(2):146–52.

2. Sly PD, Gangell CL, Chen L, Ware RS, Ranganathan S, Mott LS, et al. Risk factors for bronchiectasis in children with cystic fibrosis. N Engl J Med. 2013 May 23;368(21):1963–70.

3. Begum N, Byrnes CA, Cheney J, Cooper PJ, Fantino E, Gailer N, et al. Factors in childhood associated with lung function decline to adolescence in cystic fibrosis. J Cyst Fibros. 2022 Nov;21(6):977–83.

4. Sanders DB, Bittner RCL, Rosenfeld M, Redding GJ, Goss CH. Pulmonary exacerbations are associated with subsequent FEV _1_ decline in both adults and children with cystic fibrosis: Pulmonary Exacerbations and FEV _1_ Decline. Pediatr Pulmonol. 2011 Apr;46(4):393–400.

5. Kettle AJ, Turner R, Gangell CL, Harwood DT, Khalilova IS, Chapman AL, et al. Oxidation contributes to low glutathione in the airways of children with cystic fibrosis. Eur Respir J. 2014 Jul;44(1):122–9.

6. Kettle AJ, Chan T, Osberg I, Senthilmohan R, Chapman ALP, Mocatta TJ, et al. Myeloperoxidase and protein oxidation in the airways of young children with cystic fibrosis. Am J Respir Crit Care Med. 2004 Dec 15;170(12):1317–23.

7. Dickerhof N, Pearson JF, Hoskin TS, Berry LJ, Turner R, Sly PD, et al. Oxidative stress in early cystic fibrosis lung disease is exacerbated by airway glutathione deficiency. Free Radic Biol Med. 2017 Dec;113:236–43.

8. Dickerhof N, Turner R, Khalilova I, Fantino E, Sly PD, Kettle AJ, et al. Oxidized glutathione and uric acid as biomarkers of early cystic fibrosis lung disease. J Cyst Fibros. 2017 Mar;16(2):214–21.

9. Fantino E, Gangell CL, Hartl D, Sly PD. Airway, but not serum or urinary, levels of YKL-40 reflect inflammation in early cystic fibrosis lung disease. BMC Pulmonary Medicine. 2014 Feb 27;14(1):28.

10. Stutz MD, Gangell CL, Berry LJ, Garratt LW, Sheil B, Sly PD. Cyanide in bronchoalveolar lavage is not diagnostic for Pseudomonas aeruginosa in children with cystic fibrosis. European Respiratory Journal. 2011 Mar 1;37(3):553–8.

11. Jh R, R B, Ng M, Z B, Rg C. Systemic deficiency of glutathione in cystic fibrosis. Journal of applied physiology (Bethesda, Md : 1985) [Internet]. 1993 Dec [cited 2023 Oct 24];75(6). Available from: https://pubmed.ncbi.nlm.nih.gov/8125859/

12. Aurora P, Stocks J, Oliver C, Saunders C, Castle R, Chaziparasidis G, et al. Quality control for spirometry in preschool children with and without lung disease. Am J Respir Crit Care Med. 2004 May 15;169(10):1152–9.

13. Stahl M, Joachim C, Blessing K, Hämmerling S, Sommerburg O, Latzin P, et al. Multiple breath washout is feasible in the clinical setting and detects abnormal lung function in infants and young children with cystic fibrosis. Respiration. 2014;87(5):357–63.

14. Rabin HR, Butler SM, Wohl MEB, Geller DE, Colin AA, Schidlow DV, et al. Pulmonary exacerbations in cystic fibrosis. Pediatr Pulmonol. 2004 May;37(5):400–6.

15. Regelmann WE, Schechter MS, Wagener JS, Morgan WJ, Pasta DJ, Elkin EP, et al. Pulmonary exacerbations in cystic fibrosis: young children with characteristic signs and symptoms. Pediatr Pulmonol. 2013 Jul;48(7):649–57.

